# Forced-rate aerobic cycling enhances motor recovery in persons with chronic stroke: A randomized clinical trial

**DOI:** 10.1101/2023.08.02.23293572

**Authors:** Susan M. Linder, Andrea Bischof-Bockbrader, Sara Davidson, Yadi Li, Brittany Lapin, Tamanna Singh, John Lee, Francois Bethoux, Jay L. Alberts

## Abstract

**Background:** The potential for intensive aerobic exercise to enhance neuroplasticity post-stroke has been theorized but not systematically investigated. Our aim was to determine the effects of forced-rate aerobic exercise paired with an abbreviated session of upper extremity (UE) repetitive task practice (FE+RTP) compared to time-matched UE repetitive task practice (RTP only) on the recovery of motor function in individuals with chronic stroke.

**Methods:** A single center randomized clinical trial was conducted from April 2019 to December 2022. Sixty individuals ≥6 months following single stroke with residual UE hemiparesis aged 18-85 were recruited. Participants were randomized 1:1 to one of two time-matched groups: FE+RTP (N=30) or RTP only (N=30). Both groups completed 90-minute sessions, 3x/week for 8 weeks. The FE+RTP group underwent 45-minutes of forced-rate aerobic exercise (FE) followed by 45-min of UE RTP. The RTP only group completed 2 sequential 45-minute sessions of UE RTP. The primary UE outcomes were the Fugl-Meyer Assessment (FMA) and the Action Research Arm Test (ARAT). The Six-minute Walk Test (6MWT) assessed walking capacity.

**Results:** Sixty individuals 60.5 (±10.6) years of age and 26.0 [13.0, 69.0] months post-stroke were enrolled, and 56 completed the study. The RTP only group completed more RTP in terms of repetitions (411.8±44.4 versus 222.8±28.4, P<0.001) and time (72.7±6.7 versus 37.8±2.4 minutes, P<0.001) compared to FE+RTP. Both groups improved on the FMA (FE+RTP, 36.2±10.1 to 44.0±11.8 and RTP only, 34.4±11.0 to 41.2±13.4, P<0.001) and ARAT (FE+RTP, 32.5±16.6 to 37.7±17.9 and RTP only, 32.8±18.6 to 36.4±18.5, P<0.001). A group main effect was not observed. The FE+RTP group demonstrated greater improvements on the 6MWT (274.9±122.0 to 327.1±141.2 meters) compared to the RTP only group (285.5±160.3 to 316.9±170.0, P=0.004).

**Conclusions:** While both interventions elicited comparable improvements in UE motor recovery, the summative value of FE+RTP in improving UE and lower extremity function was demonstrated.

**Trial Registration:** ClinicalTrials.gov: NCT03819764

## Introduction

Advances in the medical management of stroke have improved survival rates by nearly one-third over the past 20 years; however, disability rates post-stroke have not changed substantially.^1^ Despite extensive resources dedicated to rehabilitation, nearly two-thirds of survivors do not regain full use of their hemiparetic upper and lower extremities resulting in the inability to use the more affected hand, deficits in locomotion and mobility, and decreased community reintegration.^2, 3^ Neuroplasticity, the ability of the central nervous system (CNS) to change in response to stimuli by strengthening existing and forming new neural connections, remains the tenet of motor recovery post-stroke.^1, 2, 4, 5^ Motor learning-based therapies to drive the recovery of upper extremity (UE) function are considered the standard of care in post-stroke rehabilitation and include constraint-induced movement therapy and repetitive task practice (RTP).^1, 5–9^ Clinical trials in the past two decades have focused on variations to these approaches of motor retraining therapies^7, 10, 11^ and titrating the optimal dosage to drive neural recovery.^8, 12^ However, the optimal method to facilitate UE motor recovery has not been determined and the causal relationship between dose and recovery remains unclear.^3, 8, 9^

Principles of motor recovery indicate that neuroplasticity is experience-dependent, requiring motor task practice to improve function. Unfortunately, the dose of therapy most individuals post-stroke receive does not meet a level of practice necessary to realize the full recovery potential.^6, 12^ There is emerging evidence to indicate that aerobic exercise (AE) improves brain function and promotes neuroplasticity.^4, 13–17^ Studies in animals and humans indicate that AE can increase levels of brain-derived neurotrophic factor (BDNF) and insulin-like growth factor-1 (IGF-1), neurotrophins thought to be key facilitators of neuroplasticity, implicated on a molecular level to encourage axonal and dendritic growth and remodeling, synaptogenesis, and synaptic transmission efficacy.^2, 4, 13, 14, 17, 18^ Numerous systematic reviews of animal and human literature have found that exercise intensity is an important factor in upregulating neurotrophins, with higher intensities eliciting greater responses compared to low-intensity AE approaches.^13, 14, 16, 17, 19^

To harness the potential neuroplastic priming effects of AE, studies conducted in animals and neurologically healthy humans indicate that AE administered immediately prior to motor task practice enhances motor skill acquisition.^18^ Our underlying hypothesis is that AE primes the CNS to facilitate neuroplasticity and its use in combination with motor task practice will optimize motor recovery.^4, 13, 15^ To evaluate this hypothesis, we conducted successful preliminary studies suggesting AE training can enhance the motor learning benefits associated with UE RTP in individuals with stroke.^20, 21^ However, a barrier is that persons post-stroke often present with considerable declines in cardiovascular fitness and endurance, which necessarily limits the duration and intensity of AE. To overcome this deconditioning, we utilized a forced-rate exercise (FE) paradigm, an approach in which the voluntary efforts of participants are supplemented to facilitate exercise of greater intensity and duration to trigger potential neuroplastic benefits.

During FE, pedaling cadence on a stationary cycle is supplemented by a motor to assist, but not replace, the voluntary efforts of the individual. We previously reported that when comparing FE to voluntary-rate exercise (VE), FE elicited greater improvements in motor recovery associated with UE RTP.^20, 21^

While our pilot studies demonstrated superiority of FE, the dose-response relationship with UE RTP was not evaluated. The aim of this project was to determine the effects of FE paired with an abbreviated session of UE RTP (FE+RTP) compared to time-matched UE RTP (RTP only) on the recovery of motor function in individuals with chronic stroke. We hypothesized that FE+RTP would yield greater improvements in UE motor recovery compared to time-matched RTP only.

## Materials and Methods

A single-center, parallel group, rater blinded randomized clinical trial with a 1:1 allocation ratio was conducted at the Cleveland Clinic between April 2019 and December 2022. The trial was funded by the National Institutes of Health (K01HD092556), registered prospectively at ClinicalTrials.gov (NCT03819764) and conducted in accordance with the Consolidated Standards of Reporting Trials (CONSORT) guidelines.

### Participants and Screening

Sixty individuals were recruited via a stroke research participant registry, clinician outreach, and support group education sessions. Inclusion criteria were: 1) ≥6 months since a single ischemic or hemorrhagic stroke; 2) UE Fugl-Meyer motor score 19-55; 3) ambulatory ≥20 meters with no more than contact guard assistance; and 4) 18-85 years of age. Exclusion criteria were: 1) hospitalization for myocardial infarction, heart failure or heart surgery within 3 months, 2) cardiac arrhythmia, 3) hypertrophic cardiomyopathy, 4) severe aortic stenosis, 5) pulmonary embolus, 6) significant contractures, 7) anti-spasticity injection to paretic UE within 3 months of enrollment and 8) other contraindications to exercise. The study was approved by the Cleveland Clinic Institutional Review Board and the informed consent process was completed with all participants.

After meeting initial screening criteria,^22^ participants underwent cardiopulmonary exercise (CPX) testing administered by an exercise physiologist at baseline and end of treatment (EOT) to determine baseline function and cardiopulmonary response to maximum exertion. Participants completed testing while on all prescribed medications to obtain an accurate measure of heart rate (HR) and blood pressure response. A 12-lead electrocardiogram was assessed prior to exercise and monitored continuously throughout exercise and recovery. An individualized ramp protocol was employed on an electronically controlled Lode cycle ergometer (Lode, Groningen, The Netherlands) using MedGraphics CardioO2/CP system (MCG Diagnostics, St. Paul, MN) with the ramp set to elicit a test duration of approximately 10 minutes. Participants were encouraged to exercise until volitional fatigue or standard test termination criteria according to the American College of Sports Medicine (ACSM) Guidelines for Exercise Testing and Prescription.^23^ Results were interpreted by the study cardiologist to determine participant safety.

### Outcomes

Clinical outcomes evaluating UE motor recovery and walking capacity were obtained at baseline, EOT, and 4 weeks following EOT (EOT+4) by the same trained physical therapist blinded to group allocation. The primary impairment-based motor outcome was the UE Fugl-Meyer Assessment (FMA)^24^, which evaluates the ability of the individual to move out of synergistic patterns toward isolated movement control.^25^ The primary functional-based motor outcome was the Action Research Arm Test (ARAT),^26^ which includes 19 tests of fine and gross motor function, grouped in categories of grasp, grip, pinch and gross movement. The Wolf Motor Function Test (WMFT) served as secondary motor outcome.^27^

The six-minute walk test (6MWT) was administered along a continuous 100-meter oval lap as a measure of walking capacity. Participants used assistive devices and orthoses as prescribed for community ambulation, with equipment remaining consistent across testing time points.

### Interventions

Following baseline testing, participants (N=60) underwent concealed randomization stratified by baseline UE FMA level (≤33 or >33) using a nonreplenished sealed opaque envelope pull method to one of two time-matched groups: 1) forced-rate aerobic exercise and repetitive task practice (FE+RTP, N=30) or 2) RTP only (N=30). Randomization was conducted by the research therapist who was not involved in the collection of outcomes. Participants in both groups completed 24 visits, 3x per week for 8 weeks, and were permitted up to 12 weeks to complete the 24 visits to allow for vacations, illness, or holidays. Study visits for both groups were matched at 90 minutes in length.

*Forced Exercise + Repetitive Task Practice:* Participants in the FE+RTP group completed a 45-minute session of supervised AE on a custom-engineered motor-assisted stationary semi-recumbent cycle ergometer, followed by 45-minutes of UE RTP. Participants were instructed to exercise within their target HR zone during the 35-minute main exercise set, which occurred between a 5-minute warm-up and 5-minute cool-down. Target HR range was individualized using the Karvonen formula at 60-80% of maximum HR based on CPX testing.^28^ Heart rate was monitored and recorded continuously using a Wahoo chest strap (Wahoo Fitness, Atlanta, GA), synced and displayed continuously on an Apple iPad (Apple, Inc, Cupertino, CA), allowing the therapist and participant to monitor aerobic intensity, facilitating adherence to prescribed values. Participants were instructed to actively pedal with the motor to achieve target HR response.

Cycling cadence was programmed by the therapist based on the individual’s ability to contribute to the workload with a target cadence of ≥75 revolutions per minute (RPM), based on results from previous studies indicating better outcomes at higher cycling cadence.^20^ Average HR during the warm-up, main set, and cool down were recorded. Blood pressure and rating of perceived exertion were recorded every 10 minutes. Training was conducted under the supervision of a physical therapist or physical therapist assistant certified in Basic Life Support. Participants completed a 45-minute session of UE RTP immediately following their FE session.

### Repetitive task practice

Repetitive task practice emphasizes highly repetitious UE tasks that are functional, goal-oriented and relevant to the individual, administered using blocked practice. The RTP approach was identical to our previous studies,^20, 21^ originally modeled after Birkenmeier and colleagues.^6, 8, 29^ Tasks were individualized to each participant’s goals and abilities and included gross and fine motor components involving proximal and distal motor control; for example, reaching to a shelf positioned at knee height, grasping a plastic cup, and placing it onto a shelf positioned at shoulder height. The RTP activities were administered by a licensed neurologic physical therapist or physical therapist assistant who tailored each task to ensure appropriate difficulty. During a 45-minute RTP session, participants completed 3-5 tasks, targeting 60-90 repetitions of each task. Time spent completing tasks and number of repetitions were recorded. The RTP intervention was administered in the same manner for both groups; however, to ensure a time-matched intervention, the FE+RTP group participated in a 45-min session immediately after 45-min of FE, while the RTP only group completed two sequential 45-min RTP sessions.

### Statistical Analysis

The power and sample size calculation was based on FMA scores from our pilot study.^21^ Assuming a minimal clinically important difference (MCID) value for the UE FMA of 4.25 points^30^ and an increase of 2*MCID for the FE+RTP group and 1*MCID for the RTP only group, an n=30 in group provided a .87 power at the 0.05 significance level to detect pairwise group differences equivalent to an effect size of .4.

Intention to treat analysis was conducted. Participant demographics, clinical characteristics, intervention parameters, and outcomes were summarized by mean with standard deviation or median with interquartile range for continuous variables, and count with percentage for categorical variables.

Mixed-effects models were used to model outcomes over time. Outcomes at baseline, EOT and EOT+4 were dependent variables. Independent variables include time point (baseline, EOT and EOT+4), group, and interaction between time and group. Subject random effect was included in the model. Interaction term was excluded if it was non-significant (p>0.05).

A responder analysis was conducted by determining the number of participants with change scores that exceed the MCID from baseline to EOT and EOT+4, for FMA, ARAT, and 6MWT. The distribution of responses for both groups were graphed using a cumulative distribution function.^31^

Statistical analyses were conducted using SAS version 9.4 (SAS Institute Inc, Cary, NC) with statistical significance established throughout at p<0.05.

## Results

Sixty participants met participation criteria, were consented, and enrolled. Participants were 60.5 (± 10.6) years of age and 58.3% male. Racial distribution included 78.3% white, 20.0% Black/African American, and 1.7% Asian. Median time since stroke was 26.0 [13.0, 69.0] months. Detailed demographics, which were similar across groups, are presented in Table 1.

**Table 1.**
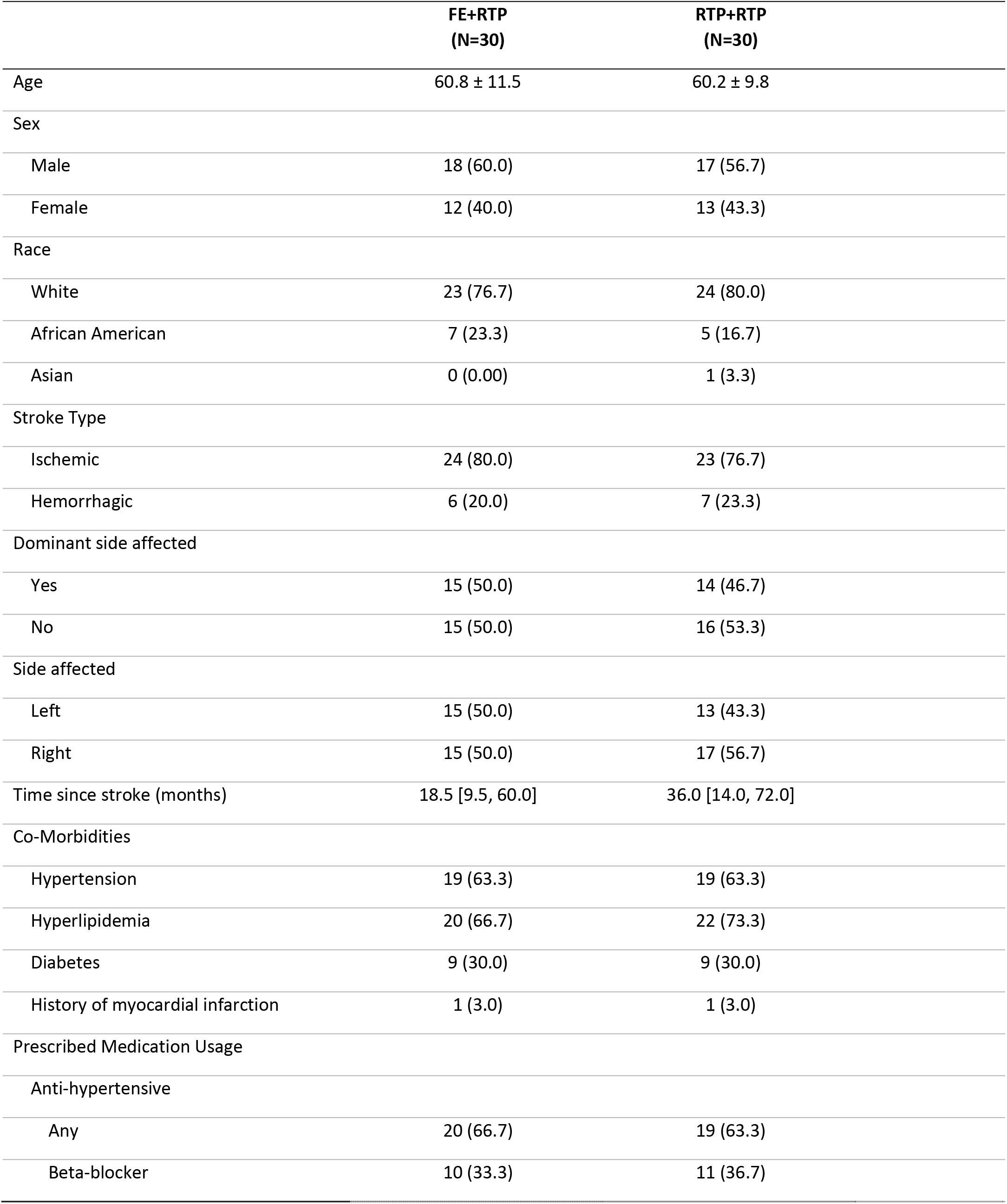
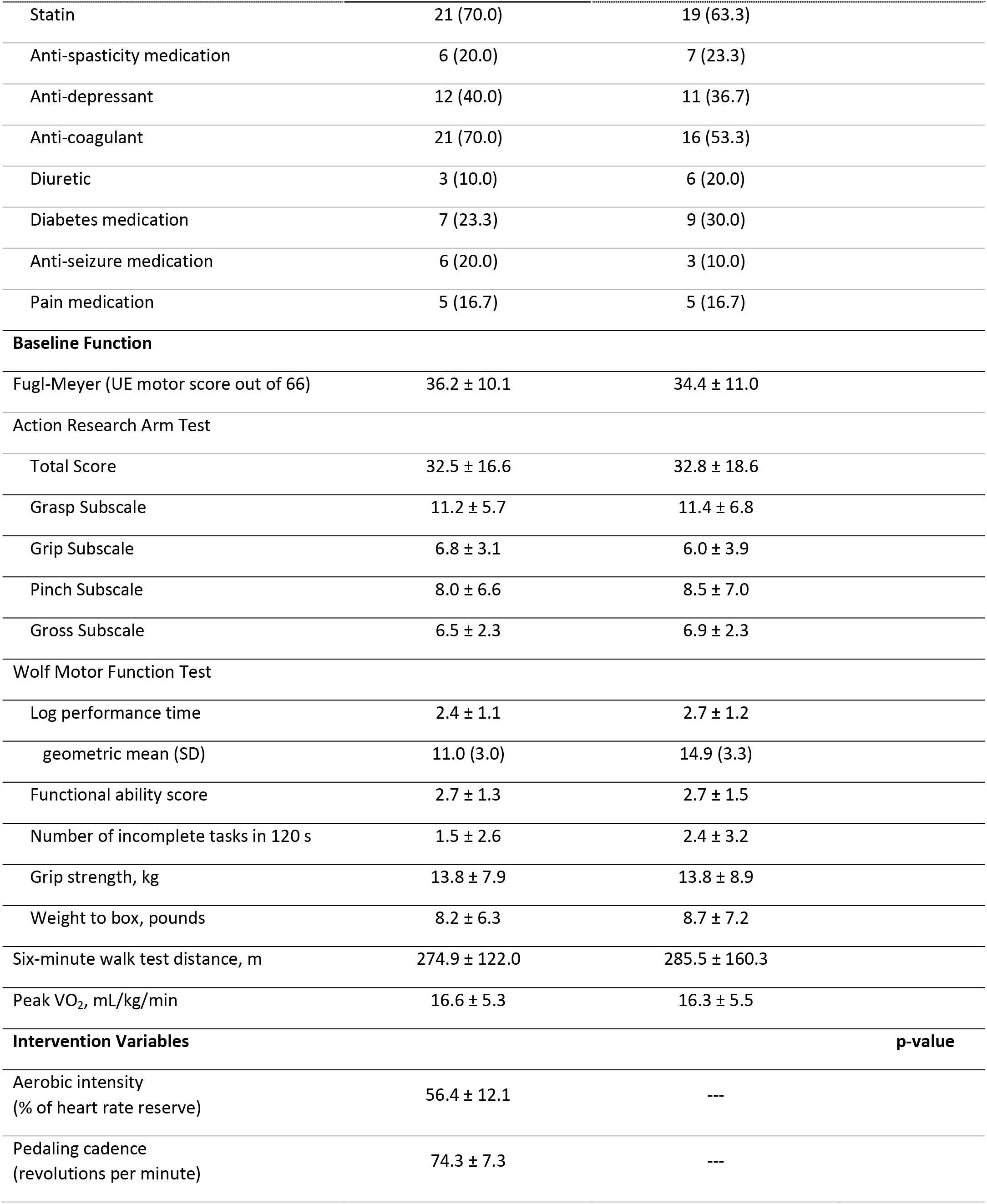
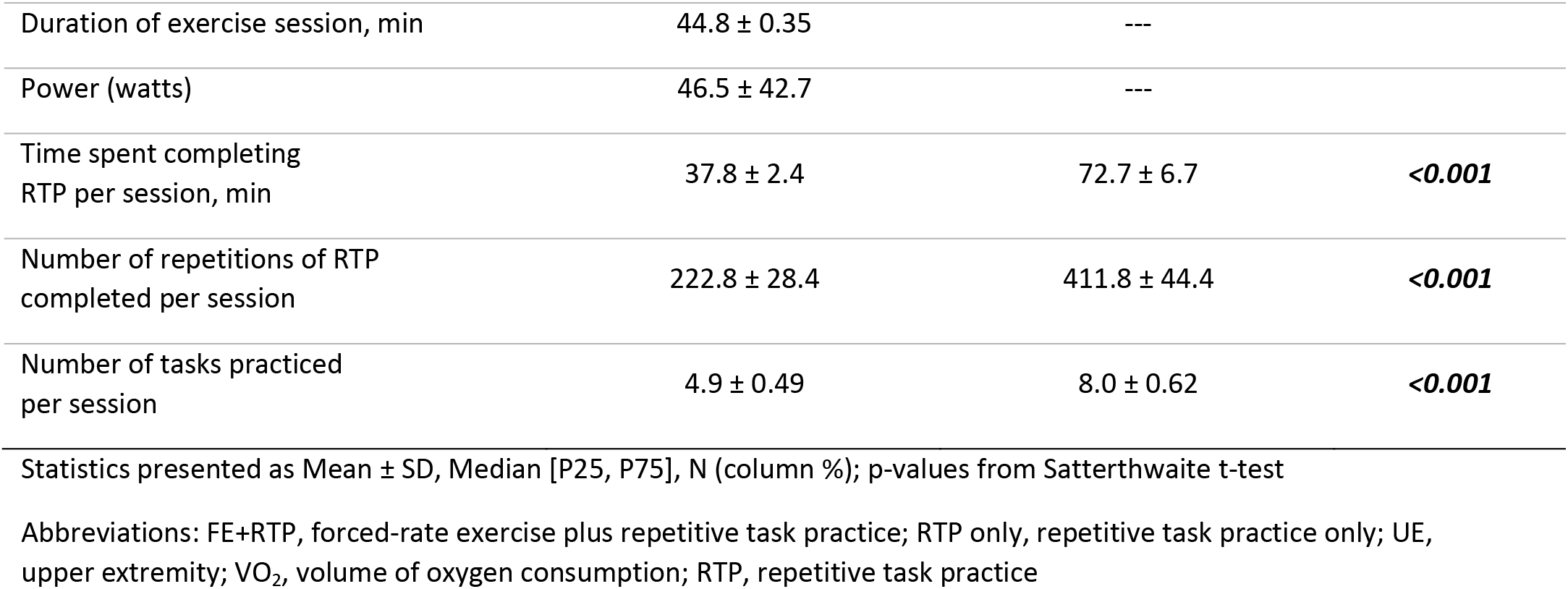
Demographics, clinical characteristics, and intervention parameters of the study sample, stratified by groups.

Both interventions were well-tolerated by participants, with an overall retention of 93.3% and no serious adverse events related to study interventions. Twenty-nine participants in the FE+RTP group and 27 in the RTP only group completed the intervention and EOT testing. Two FE+RTP participants impacted by the mandatory Covid 19 shutdown completed 20 and 21 sessions rather than the planned 24 sessions. The study team conducted EOT testing immediately prior to the shutdown to preserve outcomes from their participation. Two additional participants randomized to FE+RTP completed only 2 and 6 sessions prior to the shutdown. Both resumed the intervention 12 weeks later, restarting with session one. The CONSORT flow diagram is shown in Figure 1.

**Figure 1.**
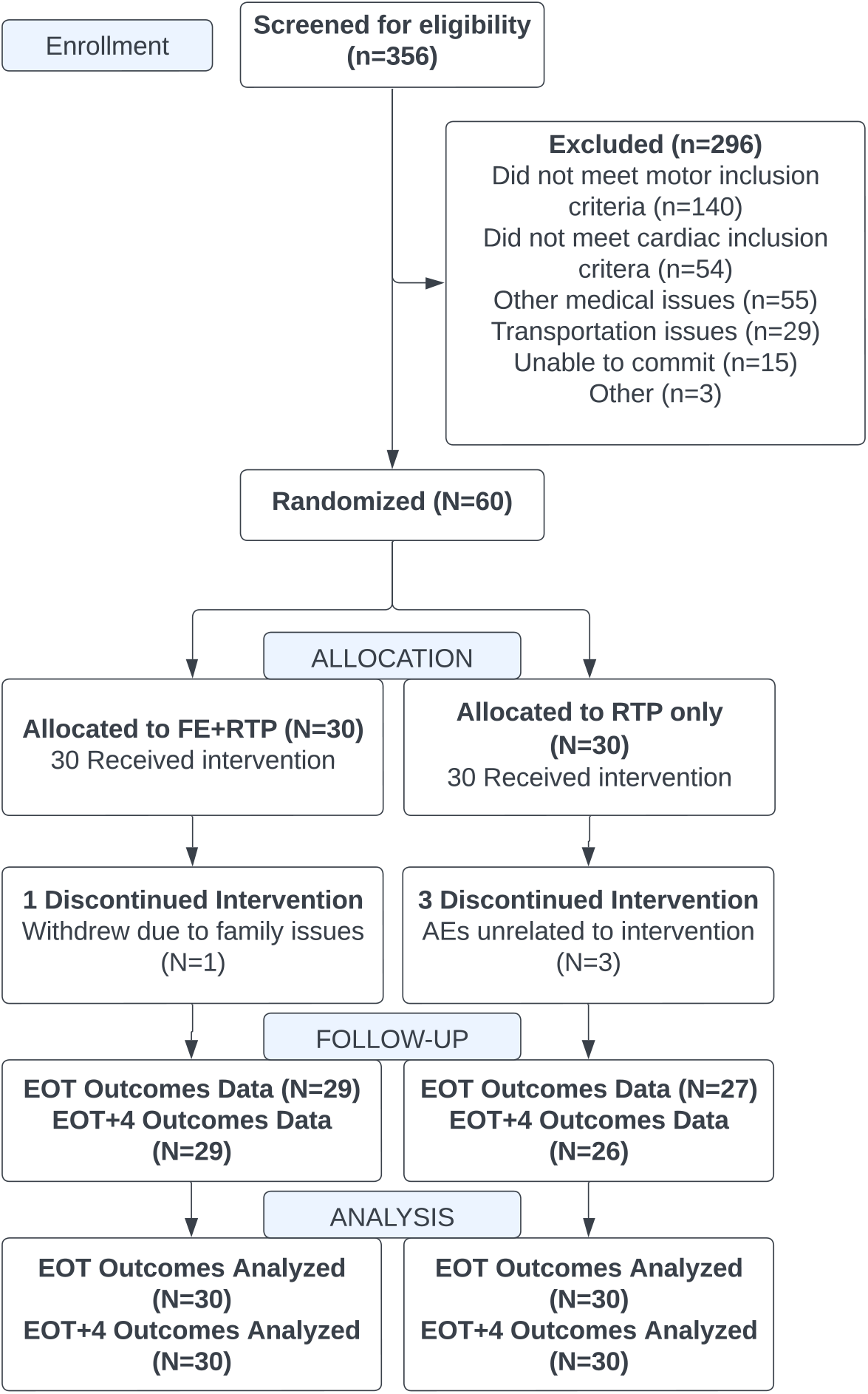
CONSORT Flow Diagram. Abbreviations: FE+RTP, forced-rate exercise plus repetitive task practice; RTP only, repetitive task practice only; AEs, adverse events; EOT, end of treatment; EOT+4, 4 weeks following end of treatment

### Intervention Variables

Average FE session duration was 44.8 (± 0.35) minutes, aerobic intensity was 56.4 (± 12.1) percent of heart rate reserve (HRR), power output was 46.5 (± 42.7) watts, and cycling cadence was 74.3 (± 7.3) RPM (Table 1). As expected, those in the RTP only group, compared to the FE+RTP group, completed nearly twice the repetitions of RTP, (411.8 ± 44.4 versus 222.8 ± 28.4, P<0.001) and spent more time completing RTP (72.7 ± 6.7 versus 37.8 ± 2.4 minutes, P<0.001).

### Upper Extremity Motor Outcomes

Significant improvements in UE motor recovery (Table 2) were measured for both groups from baseline to EOT in the UE FMA, (FE+RTP, 36.2 ± 10.1 to 44.0 ± 11.8 and RTP only, 34.4 ± 11.0 to 41.2 ± 13.4, P<0.001) and the ARAT (FE+RTP, 32.5 ± 16.6 to 37.7 ± 17.9 and RTP only, 32.8 ± 18.6 to 36.4 ± 18.5, P<0.001). A group main effect was not present for any UE outcomes (Figure 3a-b).

**Figure 2.**
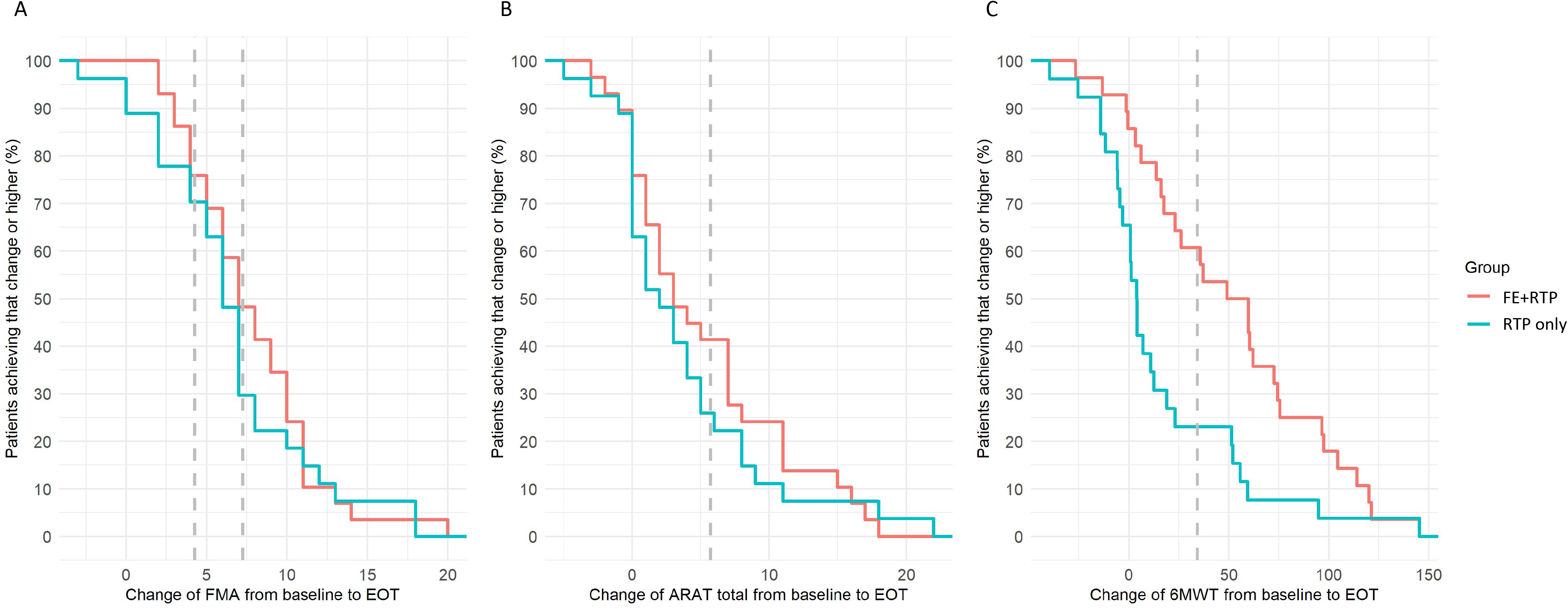
Cumulative distribution function of change for outcomes from baseline to EOT. Change scores for the Upper Extremity Fugl-Meyer Motor score (FMA, 2a), Action Research Arm Test (ARAT, 2b), and Six-minute walk test (6MWT, 2c) are shown with the minimal clinically important difference (MCID) range or value depicted by the blue horizontal shading or line, respectively. The y-axis shows the proportion of patients at each point along the outcome scale (x-axis) who experience change at that point or lower. While both groups made statistically significant improvement on the FMA and ARAT, more participants in the FE+RTP group exceeded MCID thresholds at EOT.

**Figure 3.**
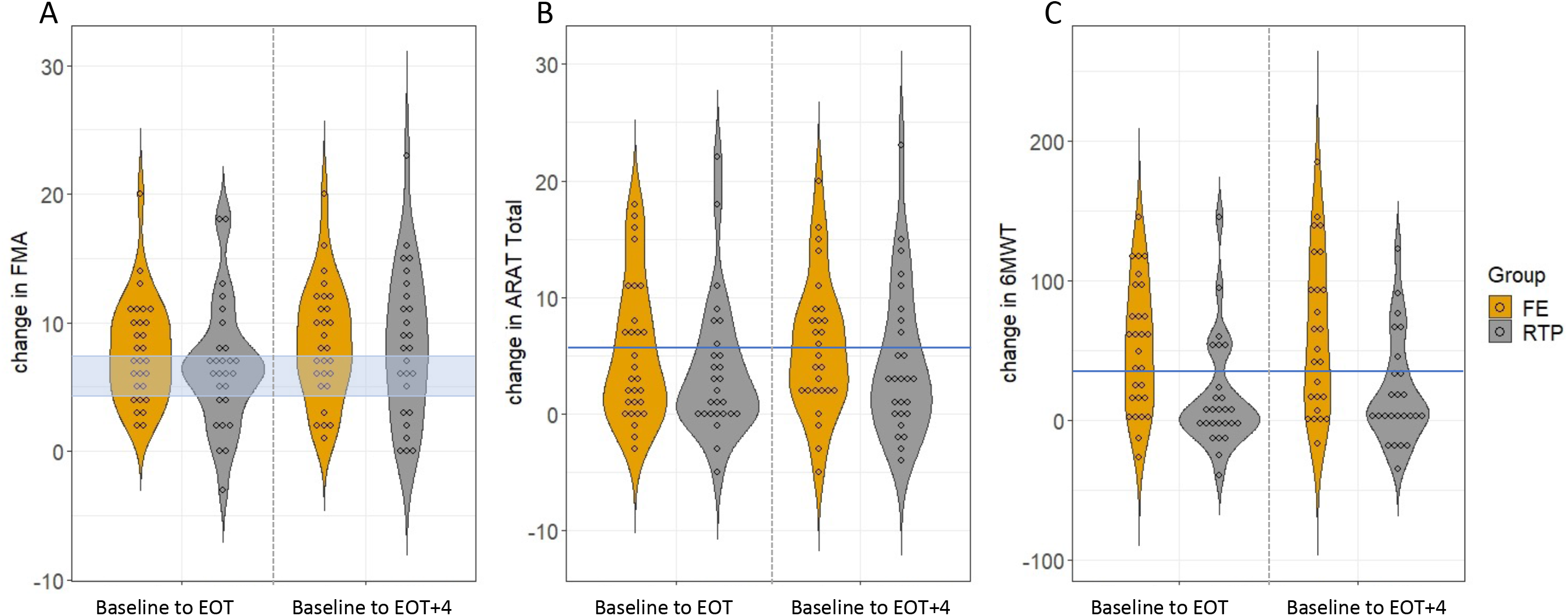
Violin dot plots depicting change scores from baseline to end of treatment (EOT, left panel) and baseline to EOT+4 weeks (right panel) with each dot representing a participant. Change scores for the Upper Extremity Fugl-Meyer Motor score (FMA, 3a), Action Research Arm Test (ARAT, 3b), and Six-minute walk test (6MWT, 3c) are shown with the minimal clinically important difference (MCID) range or value depicted by the blue horizontal shading or line, respectively. While both groups made significant improvement on the FMA and ARAT, more participants in the FE+RTP group exceeded MCID thresholds at all time points with the exception of FMA ≥7.25 at EOT+4. With respect to change in walking capacity measured by the 6MWT, only the FE+RTP group made significant improvements at both time points with group means exceeding MCID.

**Table 2.**
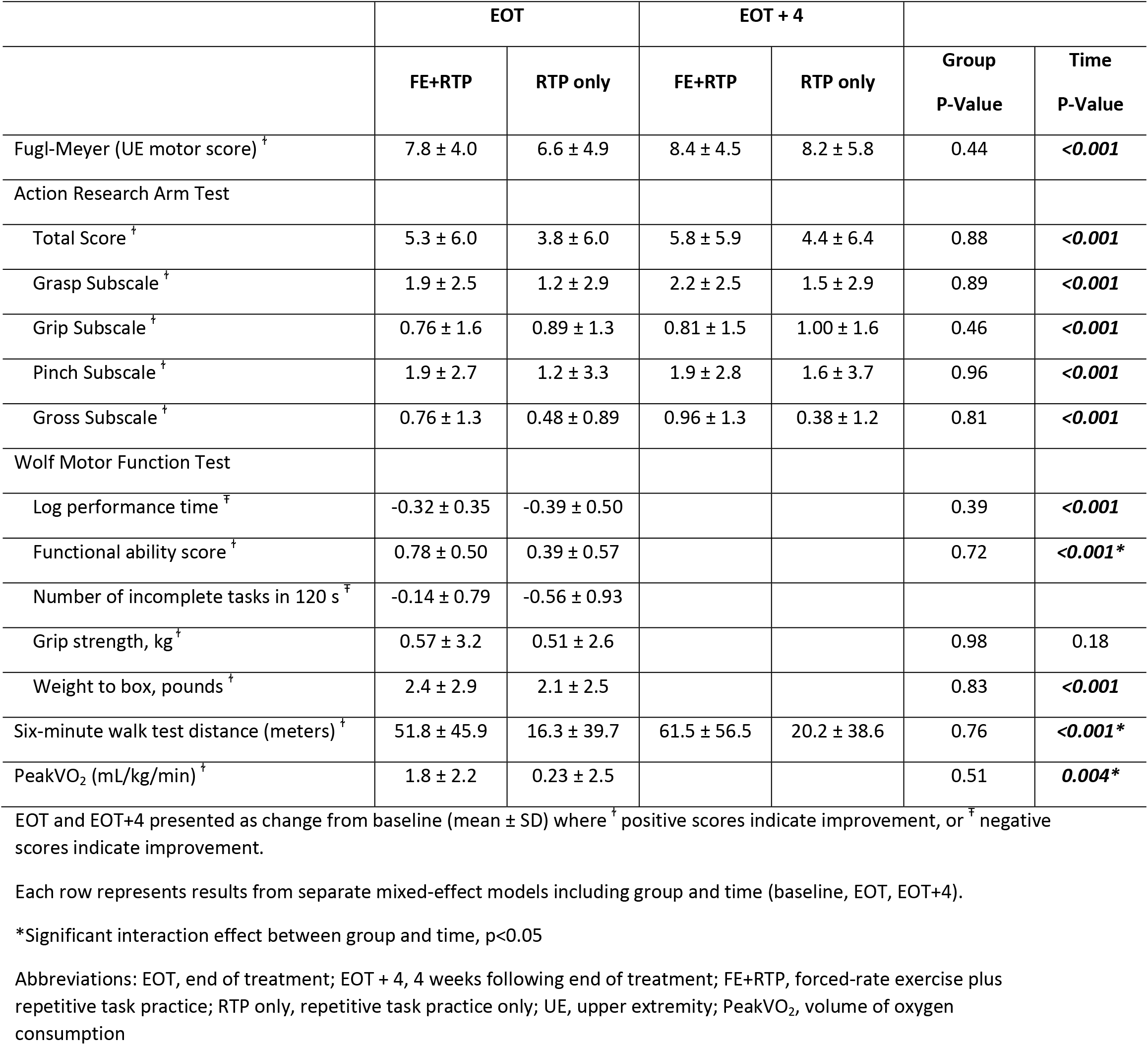
Changes from Baseline to EOT and EOT+4 by Group.

### Walking Capacity and Cardiovascular Fitness

As hypothesized, the FE+RTP group demonstrated significantly greater improvements in the 6MWT (274.9 ± 122.0 to 327.1 ± 141.2 meters) compared to the RTP only group (285.5 ± 160.3 to 316.9 ± 170.0, P=0.004, Table 2, Figure 3c) and aerobic capacity measured by peak oxygen consumption (FE+RTP, 16.6 ± 5.3 to 18.3 ± 6.4 versus RTP only, 16.3 ± 5.5 to 17.2 ± 5.6 mL/kg/min, P=0.024, Table 2).

### Responder Analysis

Change scores for the FMA, ARAT, and 6MWT that exceeded the respective MCIDs are shown in Table 3 and Figure 2. More participants in the FE+RTP group exceeded MCID thresholds at EOT and EOT+4 for all outcomes, with the exception of FMA ≥7.25 at EOT+4.

**Table 3.** Percentage of patients with change scores exceeding minimal clinically important difference.

| **Change scores exceeding MCIDs at EOT** | **Total**<br>**N (%)** | **FE+RTP**<br>**N (%)** | **RTP only**<br>**N (%)** | **Chi-square**<br>**P-value** |
| --- | --- | --- | --- | --- |
| Fugl-Meyer Assessment $\geq 4.25$ | 41 (73.2) | 22 (75.9) | 19 (70.4) | 0.64 |
| Fugl-Meyer Assessment $\geq 7.25$ | 22 (39.3) | 14 (48.3) | 8 (29.6) | 0.15 |
| Action Research Arm Test Total $\geq 5.7$ | 19 (33.9) | 12 (41.4) | 7 (25.9) | 0.22 |
| Six-minute walk test $\geq 34.4$ | 23 (42.6) | 17 (60.7) | 6 (23.1) | **0.005** |
| **Change scores exceeding MCIDs at EOT+4\*** |  |  |  |  |
| Fugl-Meyer Assessment $\geq 4.25$ | 42 (77.8) | 23 (82.1) | 19 (73.1) | 0.42 |
| Fugl-Meyer Assessment $\geq 7.25$ | 29 (53.7) | 15 (53.6) | 14 (53.9) | 0.98 |
| Action Research Arm Test Total $\geq 5.7$ | 21 (39.6) | 13 (48.2) | 8 (30.8) | 0.20 |
| Six-minute walk test $\geq 34.4$ | 21 (42.0) | 15 (60.0) | 6 (24.0) | **0.010** |

\*Subset of patients with available change scores
Abbreviations: FE+RTP, forced-rate exercise plus repetitive task practice; RTP only, repetitive task practice only; MCID, minimal clinically important difference; EOT, end of treatment; EOT+4; 4 weeks following end of treatment

Significantly more FE+RTP participants exceeded MCID on the 6MWT at EOT and EOT+4 than the RTP only group.

## Discussion

An intervention comprised of AE and RTP produced similar improvements in UE motor performance as an RTP intervention that was twice as long. Similar motor response with half the RTP dose suggests that AE may be priming the CNS to optimize neuroplasticity in persons with stroke and should be considered as an adjunct to traditional RTP. Unique to the FE+RTP group, significant and clinically meaningful improvements in locomotion and cardiovascular fitness were elicited. In sum, the FE+RTP group demonstrated greater overall gains in functional recovery than the RTP only group.

Recommendations for individuals post-stroke to engage in AE are not new; however, the benefits for CNS function have historically not been emphasized.^32^ Sustained and intensive AE has been shown to elicit vascular and neurochemical responses within the CNS, hypothesized as the mechanism by which AE facilitates neuroplasticity^4, 15, 16, 33–36^. Based on these data, our scientific premise was that intensive AE administered through FE would prime the CNS and optimize recovery associated with motor task practice. We used FE to supplement the voluntary efforts of participants to overcome barriers to completing high intensity AE that individuals post-stroke experience, including hemiparesis, fatigue, spasticity, impaired balance, and reduced cardiovascular fitness.^37^ Participants in the FE+RTP group exercised for 45 minutes at an aerobic intensity (56.4 percent of HRR) that is nearly equivalent to ACSM exercise guidelines for the general population.^38^ Therefore, supplementing the individuals’ voluntary efforts using FE facilitated a relatively intensive exercise duration (i.e.: 45 minutes/session) without compromising aerobic intensity.

Because FE involved lower limb cycling, it was unlikely to elicit a transfer of training effect to impact UE function; thus, we hypothesize that a global priming effect was induced, enhancing UE recovery. The MCID for the FMA ranges from 4.25 to 7.25 points.^30^ Despite half the RTP dosage, mean change scores for the FE+RTP group exceeded the upper value of MCID range for the FMA at both time points and responder analyses (Table 3) reveal that more participants in the FE+RTP group exceeded the MCID for the ARAT at both time points. In a previous study investigating RTP dosage, Lang and colleagues reported change in ARAT scores of 5.8 and 5.1 for participants receiving 13.6 and 20 hours of UE RTP, respectively,^8^ while our FE+RTP group demonstrated a 5.3 point change after 18 hours of RTP. By comparison, the RTP only group receiving a total of 36 hours of RTP improved by 3.8 points. We acknowledge that a gap is the investigation of mechanisms that may be responsible for the improvements observed. Future studies have been proposed that include neural and biochemical mechanisms that underlie exercise-induced plasticity.

While not designed to improve walking, a significant and meaningful by-product of FE+RTP was improvements in walking capacity, with a change in 6MWT distance of 52.2 meters, exceeding the MCID of 34.4 meters.^39^ These results approach the robust improvements in 6MWT values recently reported by Boyne and colleagues after 8 weeks of high-intensity interval treadmill-based gait training administered 3x/week^40^ and suggest that FE improves locomotor function, dispelling an assumption that cycling provides limited transfer of training to walking.^41^ An interim analysis of data from the FE+RTP group revealed normalization of gait kinematics and kinetics associated with increased gait velocity following FE, indicating participants improved locomotor control and did not exaggerate existing compensatory strategies to walk faster.^42^ Task-specificity, a fundamental concept in motor learning, has long been considered critical for neuroplasticity; yet our FE+RTP intervention did not involve task-specific gait training. While cycling and walking are different tasks, improvements in LE kinematics and kinetics from cycling may transfer to gait, as both require the rapid reciprocal activation and relaxation of lower extremity muscles synergistically, and some have proposed similar controlling neural networks.^43–46^ Pedaling data from our pilot study revealed increased hemiparetic limb torque production, improved symmetry of force production, and diminished roughness index (a measure of motion smoothness) as a result of cycling.^47, 48^ Increased somatosensory input, muscle power, lower limb motor control, and/or endurance may have elicited these gains.^49, 50^

### Conclusions and Future Directions

Our findings that an aerobic cycling intervention administered immediately prior to UE RTP can elicit robust improvements in UE function and locomotion comparable to task-specific training approaches can have significant implications on clinical practice. The FE+RTP intervention was tolerated by all participants without serious adverse events and its summative value in improving UE, lower extremity, and cardiovascular outcomes was demonstrated. Future studies employing mechanistic and clinical outcomes to elucidate the role of AE in eliciting a global response in the CNS and facilitating neuroplasticity are warranted.

## Data Availability

An application is pending to submit data from this trial to the NIH DASH database.

## Acknowledgements

The authors acknowledge Anson Rosenfeldt, DPT, and Cindy Clark, OT for administering study-related interventions.

